# High Community SARS-CoV-2 Antibody Seroprevalence in a Ski Resort Community, Blaine County, Idaho, US. Preliminary Results

**DOI:** 10.1101/2020.07.19.20157198

**Authors:** Colleen C. McLaughlin, Margaret K. Doll, Kathryn T. Morrison, William L. McLaughlin, Terry O’Connor, Anton M. Sholukh, Emily L. Bossard, Khamsone Phasouk, Emily S. Ford, Kurt Diem, Alexis M. Klock, Keith R. Jerome, Lawrence Corey

## Abstract

Community-level seroprevalence surveys are needed to determine the proportion of the population with previous SARS-CoV-2 infection, a necessary component of COVID-19 disease surveillance. In May, 2020, we conducted a cross-sectional seroprevalence study of IgG antibodies for nucleocapsid of SARS-CoV-2 among the residents of Blaine County, Idaho, a ski resort community with high COVID-19 attack rates in late March and Early April (2.9% for ages 18 and older). Participants were selected from volunteers who registered via a secure web link, using prestratification weighting to the population distribution by age and gender within each ZIP Code. Participants completed a survey reporting their demographics and symptoms; 88% of volunteers who were invited to participate completed data collection survey and had 10 ml of blood drawn. Serology was completed via the Abbott Architect SARS-CoV-2 IgG immunoassay. Primary analyses estimated seroprevalence and 95% credible intervals (CI) using a hierarchical Bayesian framework to account for diagnostic uncertainty. Stratified models were run by age, sex, ZIP Code, ethnicity, employment status, and *a priori* participant-reported COVID-19 status. Sensitivity analyses to estimate seroprevalence included base models with post-stratification for ethnicity, age, and sex, with or without adjustment for multi-participant households. IgG antibodies to the virus that causes COVID-19 were found among 22.7% (95% CI: 20.1%, 25.5%) of residents of Blaine County. Higher levels of antibodies were found among residents of the City of Ketchum 34.8% (95% CI 29.3%, 40.5%), compared to Hailey 16.8% (95%CI 13.7%, 20.3%) and Sun Valley 19.4% (95% 11.8%, 28.4%). People who self-identified as not believing they had COVID-19 had the lowest prevalence 4.8% (95% CI 2.3%, 8.2%). The range of seroprevalence after correction for potential selection bias was 21.9% to 24.2%. This study suggests more than 80% of SARS-CoV-2 infections were not reported. Although Blaine County had high levels of SARS-CoV-2 infection, the community is not yet near the herd immunity threshold.

## Introduction

Blaine County, Idaho experienced a large outbreak of COVID-19 in early March through mid-April 2020.^1^ Among 17,600 residents age 18 and older, 505 (2.9%) cases were reported through the end of June 2020. Official counts likely underestimate COVID-19 infection, particularly in a setting of early United States (U.S.) transmission, due to inadequate SARS-CoV-2 testing and strict testing criteria, bias towards detection of severe disease, and undiagnosed asymptomatic infection. Community seroprevalence surveys are needed to estimate the cumulative incidence of SARS-CoV-2 infection.

Recent COVID-19 seroprevalence studies have been methodologically scrutinized for inadequate methodology and potential selection bias.^2^ We employed best methodological practices to estimate the seroprevalence of IgG antibodies to COVID-19 in Blaine County, a high seroprevalence setting.

## Methods

### Population

Blaine County in south central Idaho has approximately 23,089 residents, and 17,611 (76.3%) residents ≥ 18 years of age.^3^ The county is home to Sun Valley Resort, a ski destination that attracts domestic and international visitors, and was likely a source of early promulgation of the county outbreak. Although the county has a large seasonal population, seasonal residents were encouraged to leave or stay away when the outbreak began. Overall, the county is approximately 77% non-Latinx white and 20% Latinx, although population demographics vary regionally.^3^ Sixty percent of county residents reside within the cities of Ketchum, Sun Valley, and Hailey.^4^

### Recruitment

From April 8–9, the City of Ketchum posted a secure website for Blaine County residents ≥ 18 years of age to volunteer for study participation. Volunteers were selected randomly for participation after stratification by ZIP Code, and by age and gender within ZIP Code. Volunteers were selected only from ZIP Codes representing Ketchum (83340), Sun Valley (83353), and Hailey (83333) due to low volunteerism in other communities. Invitations to participate were emailed to sampled volunteers with a link to an electronic consent statement and questionnaire on demographic and symptom history. A blood collection appointment was provided upon completion of consent and questionnaire. All materials were available in English or Spanish. The study was approved by the Fred Hutchinson Cancer Research Center (Fred Hutch) Institutional Review Board.

### Specimen Collection and Antibody testing

Blood was collected from May 4–19, 2020 using standard protocols into 10 cc vials with acid citrate dextrose additive to prevent clotting and shipped overnight to the Fred Hutch laboratory. Plasma was separated from cellular fraction by centrifugation at 1200 x g for 15 minutes, transferred into cryovials, and aliquots were sent to the University of Washington for testing via the Abbott Architect SARS-CoV-2 IgG chemiluminescent microparticle immunoassay, according to manufacturer protocols. In validation studies,^5–10^ assay sensitivity ranged from 92.9–100% (≥ 14 days post-symptom onset; deemed relevant based on timing of outbreak/blood collection), and specificity from 99.6–100% using pre-COVID-19 specimens. Qualitative results were shared with individual participants.

### Analytic Methods

We estimated seroprevalence and 95% credible intervals (CI) using a hierarchical Bayesian framework to account for diagnostic sensitivity and specificity.^5–11^ Stratified models were used to estimate seroprevalence by age, sex, ZIP Code, ethnicity, employment status, and *a priori* participant-reported COVID-19 status.

Additionally, we adjusted for ethnicity, age, and sex using post-stratification population weighting.^3^ Because some participants were sampled from the same household, we ran two post-stratification models where: (i) households with >1 participant were excluded, or (ii) a random sample of 1 household member from distinct households was included (run on 5 different samples). Households were identified by participant address with manual review to ensure the address represented a single-family unit.

We applied the Bayes’ odds-likelihood ratio formula to estimate seroprevalence corrected for selection bias, which may have occurred if residents differentially volunteered based on their disease history. We estimated a range for the probability of self-selection (*i.e*. likelihood ratio) by comparing the proportion of study participants who were likely to be counted in public health surveillance COVID-19 counts (see definitions in Figure 1) relative to official county cumulative incidence of COVID-19. For simplicity, we assumed volunteer behavior was the same among people who had COVID-19, irrespective of whether they were included in public health surveillance.

**Figure 1.**
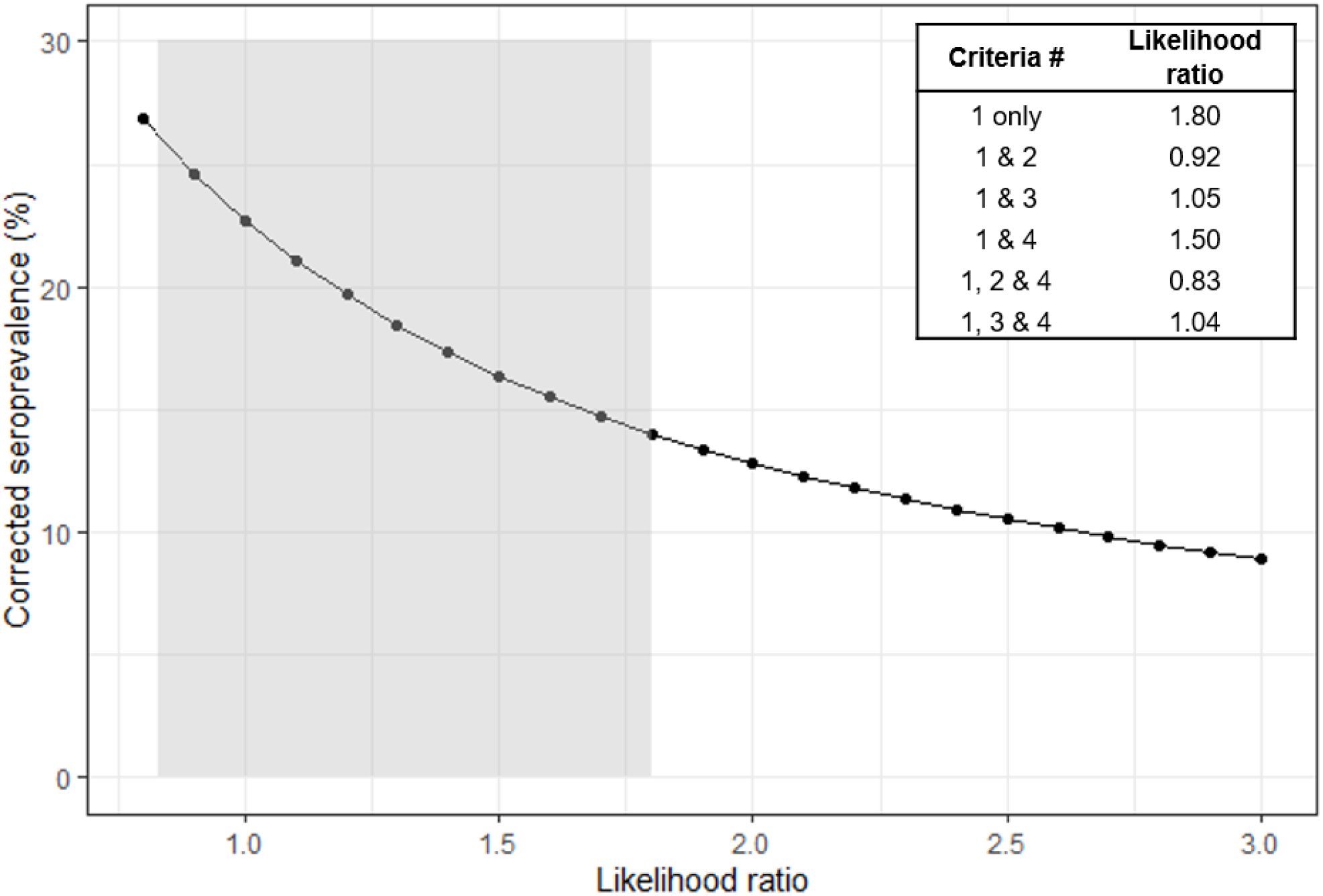
Corrected seroprevalence estimates using Bayes’ odds-likelihood formula. Caption: Plausible values for the likelihood ratio of self-selection were estimated using the following criteria in comparison with the proportion from public health surveillance data. Corrected seroprevalence values based upon these criteria are shaded in grey. \**Criteria*/*definitions used to identify a confirmed case among study participants*: 1 = participant self-report: “Yes, a medical professional told me that I had coronavirus” 2 = participant reported seeking medical care for symptoms 3 = participant reported hospitalization for symptoms 4 = participant was IgG positive for SARS-CoV-2

## Results

A total of 2,655 Blaine County residents volunteered for the study. Email invitations were sent to 1,033 volunteers. The enrollment survey was completed by 972 (94.1%) people; 917 (88.8%) also completed a blood draw. We identified 785 distinct households; 663 (68%) households had a single participant.

Of 917 participants, 208 (22.7%, 95% CI: 20.0%, 25.5%) had IgG antibodies. This estimate did not change meaningfully with adjustment for diagnostic validity, post-stratification population weighting, and multi-participant households (Table 1).

**Table 1.** Seroprevalence and 95% credible intervals (95% CI) of IgG antibodies to COVID-19 estimated by various models, Blaine County Idaho, April 2020.

| **Overall Seroprevalence** | **# Participants** | **% Positive** | **95% CI** |
| --- | --- | --- | --- |
| **Crude** | 917 | 22.7% | (20.0%, 25.5%) <sup>a</sup> |
| **Model: Diagnostic uncertainty** | 917 | 22.7% | (20.1%, 25.5%) |
| **Model: Diagnostic uncertainty + post-stratification <sup>b,c</sup>** | 917 eligible;<br>774 no missing data | 23.1% | (19.8%, 26.5%) |
| **Model: Diagnostic uncertainty + post-stratification <sup>b,c</sup> + households (excludes participants from households with multiple participants)** | 663 eligible;<br>562 no missing data | 23.4% | (19.6%, 27.5%) |
| **Model: Diagnostic uncertainty + post-stratification <sup>b,c</sup> + households (random sample of 1 participant from households with multiple participants)** | 785 eligible;<br># participants variable<br>(sample #1 run = 663<br>no missing data) | 22.9% | (19.4%, 26.6%) |

<sup>a</sup>Exact binomial confidence intervals;
<sup>b</sup>External population = SEER County-Level Population Files – 19 Age groups” reference data for Blaine County 2018 (https://seer.cancer.gov/popdata/download.html);
<sup>c</sup>Post-stratification for age (4 cats: 18-29, 30-49, 50-64, 65+ years), sex, ethnicity

Highest seroprevalence was observed in Ketchum, with seroprevalence in Sun Valley and Hailey below 20% (Table 2). Seroprevalence varied by self-reported COVID-19 status, ranging from 83.6% (95% CI: 72.0%, 92.8%) among participants reporting a medical diagnosis to 4.8% (95% CI: 2.3%, 8.2%) among those who did not believe they had COVID-19. Participants 18–29 years of age had the highest seroprevalence; older age groups showed little variation.

**Table 2.**
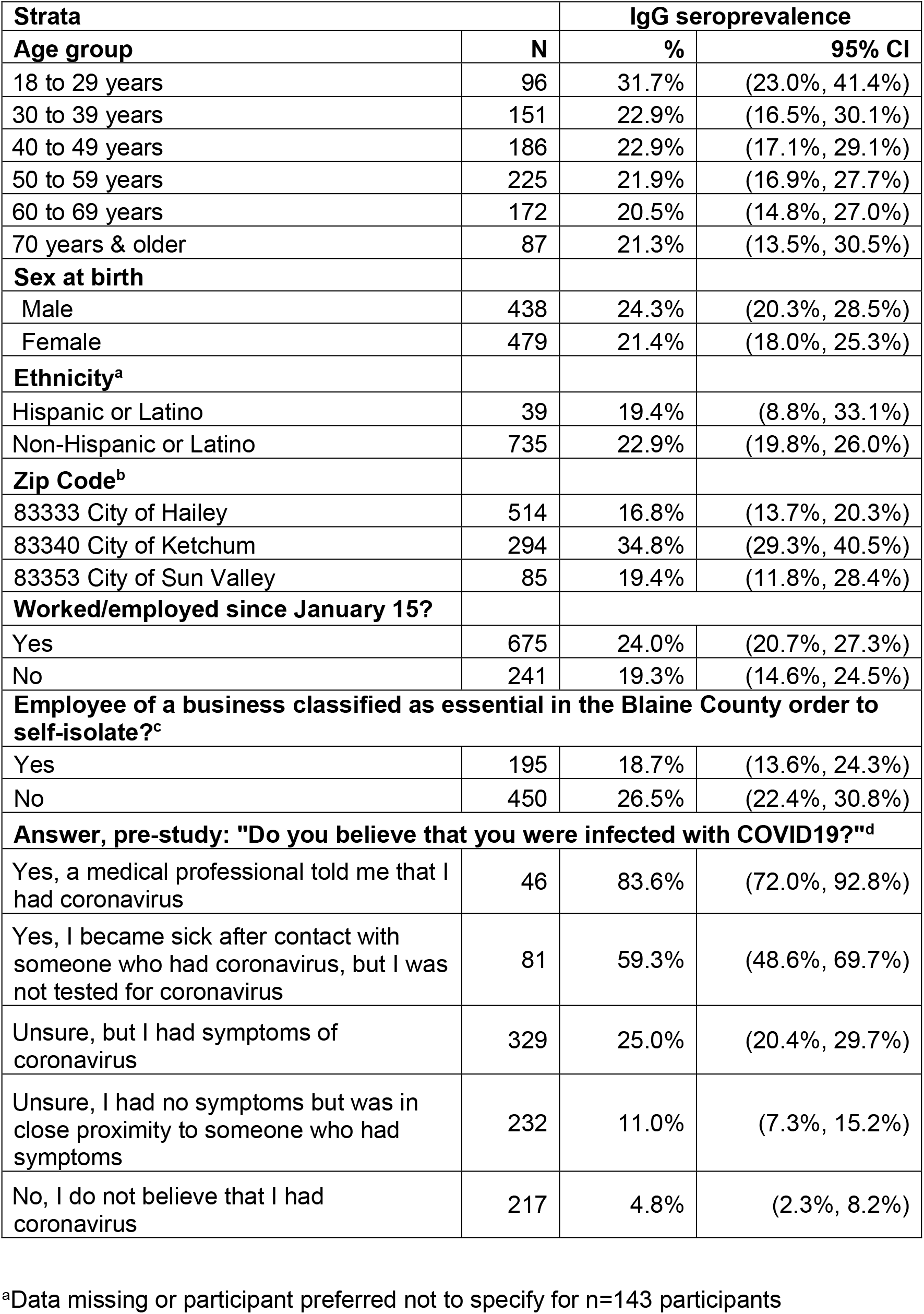
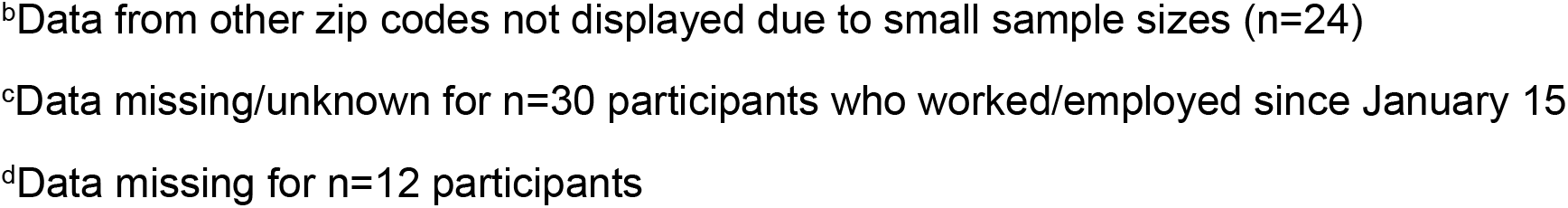
Stratified estimates and 95% credible intervals (95% CI) of seroprevalence after adjustment for diagnostic uncertainty, Blaine County Idaho, April 2020.

| **Strata** |  | **IgG seroprevalence** |  |
| --- | --- | --- | --- |
| **Age group** | **N** | **%** | **95% CI** |
| 18 to 29 years | 96 | 31.7% | (23.0%, 41.4%) |
| 30 to 39 years | 151 | 22.9% | (16.5%, 30.1%) |
| 40 to 49 years | 186 | 22.9% | (17.1%, 29.1%) |
| 50 to 59 years | 225 | 21.9% | (16.9%, 27.7%) |
| 60 to 69 years | 172 | 20.5% | (14.8%, 27.0%) |
| 70 years & older | 87 | 21.3% | (13.5%, 30.5%) |
| **Sex at birth** |  |  |  |
| Male | 438 | 24.3% | (20.3%, 28.5%) |
| Female | 479 | 21.4% | (18.0%, 25.3%) |
| **Ethnicity<sup>a</sup>** |  |  |  |
| Hispanic or Latino | 39 | 19.4% | (8.8%, 33.1%) |
| Non-Hispanic or Latino | 735 | 22.9% | (19.8%, 26.0%) |
| **Zip Code<sup>b</sup>** |  |  |  |
| 83333 City of Hailey | 514 | 16.8% | (13.7%, 20.3%) |
| 83340 City of Ketchum | 294 | 34.8% | (29.3%, 40.5%) |
| 83353 City of Sun Valley | 85 | 19.4% | (11.8%, 28.4%) |
| **Worked/employed since January 15?** |  |  |  |
| Yes | 675 | 24.0% | (20.7%, 27.3%) |
| No | 241 | 19.3% | (14.6%, 24.5%) |
| **Employee of a business classified as essential in the Blaine County order to self-isolate?<sup>c</sup>** |  |  |  |
| Yes | 195 | 18.7% | (13.6%, 24.3%) |
| No | 450 | 26.5% | (22.4%, 30.8%) |
| **Answer, pre-study: "Do you believe that you were infected with COVID19?"<sup>d</sup>** |  |  |  |
| Yes, a medical professional told me that I had coronavirus | 46 | 83.6% | (72.0%, 92.8%) |
| Yes, I became sick after contact with someone who had coronavirus, but I was not tested for coronavirus | 81 | 59.3% | (48.6%, 69.7%) |
| Unsure, but I had symptoms of coronavirus | 329 | 25.0% | (20.4%, 29.7%) |
| Unsure, I had no symptoms but was in close proximity to someone who had symptoms | 232 | 11.0% | (7.3%, 15.2%) |
| No, I do not believe that I had coronavirus | 217 | 4.8% | (2.3%, 8.2%) |

<sup>a</sup>Data missing or participant preferred not to specify for n=143 participants
<sup>b</sup>Data from other zip codes not displayed due to small sample sizes (n=24)
<sup>c</sup>Data missing/unknown for n=30 participants who worked/employed since January 15
<sup>d</sup>Data missing for n=12 participants

Using various criteria, we estimated that participants who were likely included in public health surveillance were 0.83 to 1.80 times more likely to participate in our study. Given this likelihood ratio range, we extrapolated a corrected seroprevalence between 14.0% to 26.1% (Figure 1), depending on the criteria used. If we assumed participants were more likely to be included in public health surveillance if they reported a medical diagnosis and were seen by a medical provider for their symptoms, then the likelihood ratio narrowed to 0.92 to 1.05, with a corrected seroprevalence between 21.9% to 24.2%.

## Discussion

To date, there are few peer-reviewed reports of community-based seroprevalence, although some municipalities have released surveillance data directly to the public. We estimate seroprevalence in Blaine County of 23%, among the highest reported by any municipality. Despite high seroprevalence, our estimates were approximately 45% lower than estimated levels required for herd immunity (67%),^12^ presuming IgG antibodies to SARS-CoV-2 nucleocapsid represent an accurate immune marker.

Extrapolating our seroprevalence to residents of Hailey, Ketchum, and Sun Valley, we estimate approximately 2500 adults had COVID-19 infection prior to May 19, indicating that less than 20% of infections were included in official case counts. The underestimate would be larger if mild or asymptomatic infections did not seroconvert or recent (7-14 day) infection.^13^ The rate of COVID-19 disease (2.9%) was 10% of the cumulative incidence found with antibody testing, which is similar to other recent U.S. studies with underestimates in official data ranging from 6% to 20%.^14,15^ The small number of county deaths (n=5) make estimating infection fatality rate unreliable.

U.S. community seroprevalence is difficult to estimate due to lack of population registers. We used pre-stratification to mitigate potential selection bias in our volunteer-based sample. We also corrected our estimates using varying self-selection probabilities by disease status. Our most conservative estimate of seroprevalence is 14% to 26%. Our best estimate, however, is 22% to 24%, based on the assumption that participants who were informed they had COVID-19 by their provider and also saw a medical provider for their symptoms were more likely to be included in public health surveillance counts.

In conclusion, we report seroprevalence among Blaine County residents between 22% to 24% using a methodologically rigorous design. Despite high, early COVID-19 transmission, this falls below herd immunity levels. Analyses are underway to examine the seroepidemiology of the outbreak.

## Data Availability

Data are not currently available

## REFERENCES

1. Coronavirus Weekly Dashboard. 2020; https://www.phd5.idaho.gov/coronavirus-dashboard/. Accessed June 26, 2020.

2. Bennett ST, Steyvers M. Estimating COVID-19 Antibody Seroprevalence in Santa Clara County, California. A re-analysis of Bendavid et al. medRxiv. 2020.

3. U.S. Population Data - 1969-2018, Released December 2019. https://seer.cancer.gov/popdata/. Accessed June 16, 2020.

4. Annual Estimates of the Resident Population for Incorporated Places: April 1, 2010 to July 1, 2019. https://www.census.gov/data/tables/time-series/demo/popest/2010s-total-cities-and-towns.html. Accessed June 16, 2020.

5. Abbott. Clinical Performance: Abbott ARCHITECT SARS-CoV-2 IgG Instructions for Use. 2020; https://www.corelaboratory.abbott/us/en/offerings/segments/infectious-disease/sars-cov-2. Accessed June 18, 2020.

6. Bryan A, Pepper G, Wener MH, et al. Performance characteristics of the Abbott Architect SARS-CoV-2 IgG assay and seroprevalence in Boise, Idaho. Journal of Clinical Microbiology. 2020.

7. Ng D, Goldgof G, Shy B, et al. SARS-CoV-2 seroprevalence and neutralizing activity in donor and patient blood from the San Francisco Bay Area. medRxiv. 2020.

8. Lu S, Paiva KJ, Grisson RD, et al. Validation and Performance Comparison of Three SARS-CoV-2 Antibody Assays. bioRxiv. 2020.

9. Tang MS, Hock KG, Logsdon NM, et al. Clinical Performance of Two SARS-CoV-2 Serologic Assays. Clin Chem. 2020.

10. Theel ES, Harring J, Hilgart H, Granger D. Performance Characteristics of Four High-Throughput Immunoassays for Detection of IgG Antibodies against SARS-CoV-2. Journal of Clinical Microbiology. 2020:JCM.01243-01220.

11. Gelman A, Carpenter B. Bayesian analysis of tests with unknown specificity and sensitivity. medRxiv. 2020.

12. Randolph HE, Barreiro LB. Herd Immunity: Understanding COVID-19. Immunity. 2020;52(5):737–741.

13. Gallais F, Velay A, Wendling M-J, et al. Intrafamilial Exposure to SARS-CoV-2 Induces Cellular Immune Response without Seroconversion. medRxiv. 2020:2020.2006.2021.20132449.

14. Silverman JD, Hupert N, Washburne AD. Using influenza surveillance networks to estimate state-specific prevalence of SARS-CoV-2 in the United States. Science translational medicine. 2020.

15. Havers FP, Reed C, Lim TW, et al. Seroprevalence of Antibodies to SARS-CoV-2 in Six Sites in the United States, March 23-May 3, 2020. medRxiv. 2020:2020.2006.2025.20140384.

